# User testing on Foods with Function Claims labelling in Japan : An attempt to establish an integrated evaluation system for the usefulness of health information materials

**DOI:** 10.1101/2024.04.23.24306252

**Authors:** Michiko Yamamoto, Ken Yamamoto, Hiromi Takano-Ohmuro, Rain Yamamoto, Junji Saruwatari

**Affiliations:** Graduate School of Pharmaceutical Sciences, Kumamoto University, 5-1 Oehonmachi, Chuo-ku, Kumamoto-city, Kumamoto, Japan; Showa Pharmaceutical University, 3-3165 Higashi-Tamagawagakuen, Machida-city, Tokyo, Japan; Faculty of Pharmacy, Musashino University, 1-1-20 Shin-machi, Nishitokyo-city, Tokyo, Japan; Faculty of Pharmacy, Keio University, 1-5-30 Shibakoen, Minato-ku, Tokyo, Japan

## Abstract

The saturation of self-care products in the market is coupled with inadequate information on their safe usage. In Japan, although foods with function claims (FFC) are prevalent, their labelling falls short in quality and effectiveness as health information, impeding consumer comprehension and proper utilization. Hence, it is imperative to establish a system that assesses the efficacy of labelling information from both provider and user perspectives. From providers’ or healthcare professionals’ perspective, we already developed a Communication Index to assess FFC labelling, which we utilized to evaluate five FFC products. Those products achieved a proficiency level of approximately 70%, falling below the acceptance criteria. Particularly, challenges were identified in understanding some of the terms and locating important information on the labels. In this study, we conducted user-testing from the user perspective for five same FFC labels to evaluate them using semi-structured interviews with 50 participants of diverse ages and sexes. A passing criterion for comprehension was set as ≥90% correct responses to all questions. Of the five FFC products, one passed the user-testing criterion with a 2-min response time; however, none passed the 1-min response time test. The proportions of correct answers were notably low for questions on diet and allergies (each 50-90%), concomitant medications (50-100%), storage (30-100%), and handling (30-100%). Participants’ comments revealed a lack of familiarity with FFC, highlighting that the terms and text in the labelling were confusing and overly technical. User-testing provides valuable insights for improving FFC labelling, thereby ensuring safe and appropriate use by aligning with consumers’ understanding and perceptions. We assessed FFC label information from both the provider and user perspectives, but neither yielded satisfactory results. Consequently, the implementation of an integrated system capable of evaluating FCC labels as health information material from both perspectives would be necessary.

## INTRODUCTION

### Health information provision and consumer understanding in Japan

Numerous self-care products, including health food items, saturate the Japanese market. However, the prevalence of inaccurate and unreliable health information sources can mislead consumers, potentially leading to inappropriate use of the product and associated health risks [1]. Health information materials serve as crucial tools for effective risk communication.

An online survey in 2016 revealed that only 16% of consumers clearly understood the characteristics of foods with function claims (FFC) [2]. Another survey in 2017 reported that 17% of consumers using health food products experienced poor physical conditions [3]. In March 2024, tragically, five people were fatally poisoned by the FFC containing the beni kōji fermented rice in Japan [4]. While the incident was likely caused by a contaminant, it underscored the challenge consumers face in checking the safety of FFC, which are readily available. Consequently, the provision of easy-to-understand information is crucial to ensuring safe product usage and empowering consumers to make informed choices. A comparative analysis of Japanese and European consumer health literacy surveys [5,6] indicated that 41.8% of Japanese respondents, 36.2% in Europe, and 30.1% in the Netherlands had difficulty understanding information on food packages [5,6].

Given the health literacy gap between professionals and consumers, establishing a communication system ensuring that the quality of information aligns with consumer needs, is imperative.

Previous studies investigated consumers’ comprehension of the nutrition facts label, health claims, and food labels using online surveys including questionnaires[7–9]. A qualitative study was conducted to investigate how claims can affect consumers’ perceptions and behavior [10]. While these studies investigated food labelling, they were not specific to FFC labelling. Although surveys have been conducted in Japan on consumers’ awareness and attitudes towards FFC [11], no surveys have been conducted to assess providers’ and consumers’ perspectives on labelling.

Currently, there is no system available in Japan for evaluating the usability of health information materials. To enhance the utility of these materials, it is vital to evaluate information from the providers’ perspective and further verify it from the users’ perspective. Previously, we developed a usefulness evaluation index for FFC labelling from the providers’ or healthcare professionals’ perspective [12]. In this study, we developed and evaluated a user test to gauge the accessibility and comprehensibility of the same FFC materials from consumers’ perspectives. In addition to user testing, we conducted interviews with a qualitative analysis of the comments obtained from the consumers. The development of these integrated methods considering the provider and consumer perspectives represents the first study on the comprehension of health information, using FFC labelling.

### Labelling of foods with health claims in Japan

Based on the Health Promotion Law, the ‘Foods with Health Claims’ system was established in April 2015 to facilitate the appropriate use of such foods for self-care [13,14]. This system comprises Foods for Specified Health Uses (FOSHU), Foods with Nutrient Function Claims, and FFC (S1 Fig). FOSHU undergoes individual reviews for efficacy and safety, and is approved by the Secretary General of the Consumer Affairs Agency (CAA) [15]. In contrast, FFC can display function claims based on scientific evidence, with the responsibility lying on the food business operator. Prior to marketing, information supporting the safety and efficacy of the product is submitted to the Secretary-General of the CAA [16].

As of 4 October 2023, there were 7,538 notified FFC [17], while 1,054 FOSHU products received approval [18]. The FFC must feature 16 specified items (Cabinet Office Ordinance No. 10, 2015; Table 1, Fig 1). Moreover, FFC labels should bear the following: the product’s name, storage method, best before date or expiration date, ingredients, additives, nutritional ingredients, total weight, calorific value of nutritional ingredients, and the name and address of the food business operator [19]. In the actual labels of the product containers and packaging, the order, font size and position of these items differ from the examples given by the CAA (Fig 1).

**Table 1.** Labelling on containers and packaging of foods with functional claims.

|  |
| --- |
| 1) A statement indicating that the product is a food with function claims |
| 2) The active ingredient with validated functionality along with the functionality of the ingredient or the food containing it |
| 3) Quantity and calorific value of the nutritional ingredient |
| 4) Quantity of the active ingredient within each recommended daily allowance |
| 5) Approximate daily allowance |
| 6) Notification number |
| 7) Contact details of the food business operator |
| 8) A statement indicating that the product has not undergone evaluation for functionality and safety by the regulatory agency |
| 9) Instructions for the mode of intake |
| 10) Cautionary information for intake |
| 11) Language promoting a well-balanced diet |
| 12) A statement outlining special precautions, if any, required for the cooking or preservation method |
| 13) A statement indicating that the product is not intended for the diagnosis, treatment, or prevention of diseases |
| 14) Information aimed at individuals with diseases, minors, pregnant or nursing women (including those planning to conceive), and lactating women |
| 15) A statement recommending individuals with diseases to consult a physician, and those taking medications to consult either a physician or a pharmacist before using the product |
| 16) A directive to discontinue product intake immediately and consult a physician in case of any physical discomfort |

**Fig 1.**
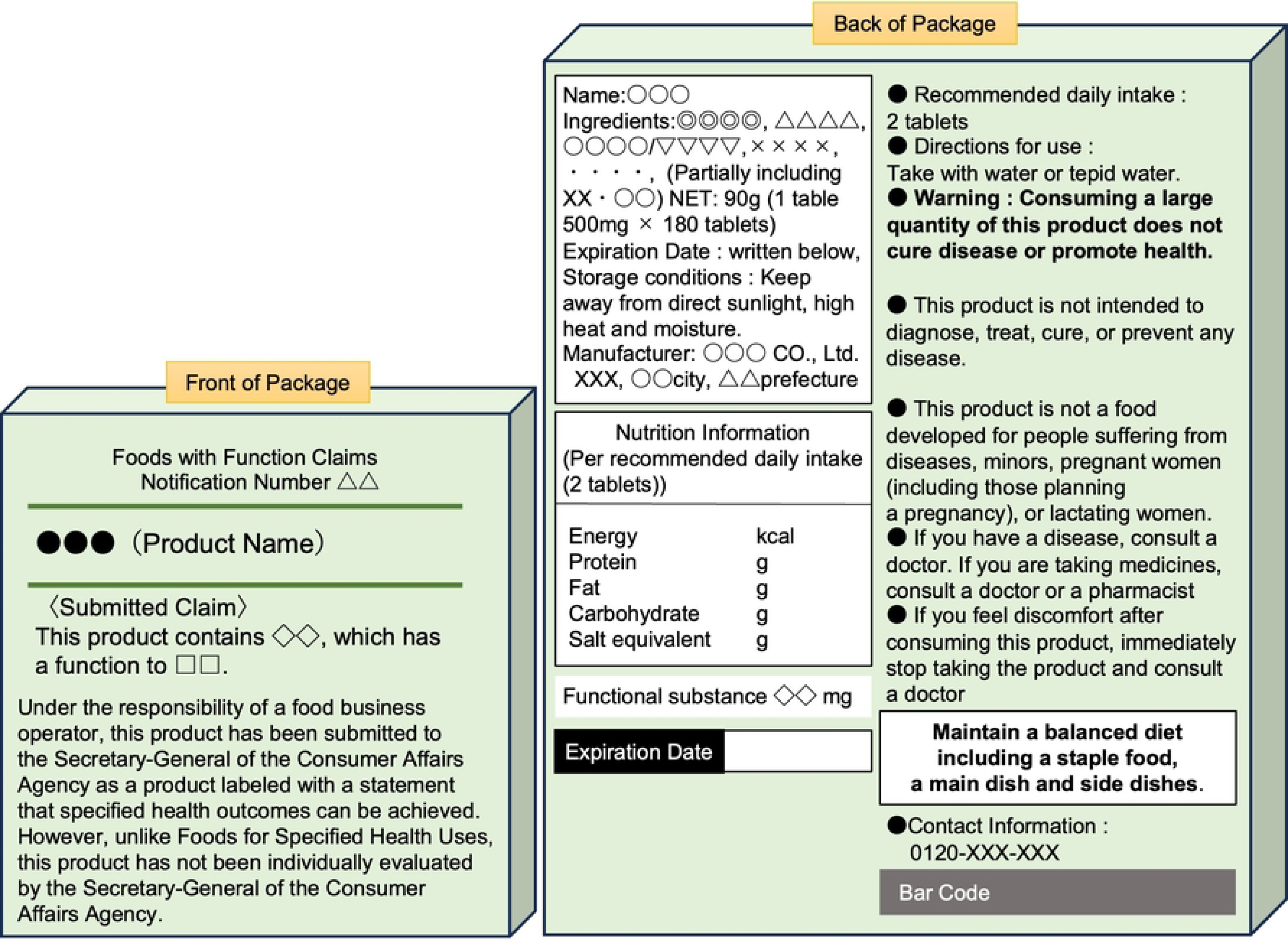
An example of the label for Foods with Function Claims.

The 16 items indicated in the Food Labelling Standards (Cabinet Office Ordinance No. 10, 2015) [20]

## Evaluation of FFC labelling

### Evaluation from the providers’ or healthcare professionals’ perspective

Foods with Health Claims should present clear information on its label, so that the information is easily understandable to consumers with diverse levels of health literacy. In recent years, public organizations in Europe and the United States have introduced standards to facilitate the creation and provision of health information that is easily understandable for consumers and patients. In the United States, various tools such as ‘Clear & Simple’ [21] and ‘Toolkit for Making Clear and Effective Information’ [22] are available. Notably, the Centers for Disease Control and Prevention (CDC) released the ‘Clear Communication Index (CCI)’ in 2014 as a research-based tool for developing and assessing public communication materials [23]. The CCI comprises 20 items, including the main message and action recommendations, with the CDC recommending a score of 90% (18 items) or higher. In this context, a group comprising six university employees, all of whom were qualified as pharmacists and public health professionals, has developed our own CCI for evaluating the FFC labelling (F-CCI) (Table 2).

**Table 2.** Evaluation index for labelling foods with function claims.

|  |  |  |
| --- | --- | --- |
| **Part A (Core)** | F-1 | Does the material contain one main message statement? (Does not necessarily have to match the Submitted Claim) |
|  | F-2 | Is the main message at the top or beginning, or on the front of the material? |
|  | F-3 | Is the main message emphasized with visual cues? |
|  | F-4 | Does the material contain at least one visual that conveys or supports the main message? |
|  | F-5 | Does the material include one or more calls to action for the primary audience? |
|  | F-6 | Do both the main message and the call to action use the active voice? |
|  | F-7 | Does the material always use words the primary audience uses? |
|  | F-8 | Does the material use bulleted or numbered lists? |
|  | F-9 | Is the material organized in chunks with headings? |
|  | F-10 | Is the most important information that the primary audience needs summarized in the first paragraph or section? |
| **Part B (Behavioral Recommendation)** | F-11 | Does the material include one or more behavioral recommendations on functionality for the primary audience? |
|  | F-12 | Does the material include one or more behavioral recommendations on safety for the primary audience? |
|  | F-13 | Does the material explain why the behavioral recommendation(s) on safety is necessary for the primary audience? |
|  | F-14 | Does the behavioral recommendation(s) include specific directions about how to perform the behavior? |
| **Part C (Numbers)** | F-15 | Does the material always present numbers that the primary audience uses? |
|  | F-16 | Does the material always explain what the numbers and units mean? |
|  | F-17 | Does the audience have to conduct mathematical calculations? |
| **Part D (Risk)** | F-18 | Does the material explain the nature (e.g., about specific harms) of the risk? |

Using the F-CCI index, we evaluated five FFC products from the perspective of healthcare professionals, achieving a level of approximately 70% (12–14 items) which we have already published [12]. None of the five products met the acceptance criteria for the following questions: ‘Does the material consistently use language familiar to the primary audience? (F-CCI Q7)’, ‘Is the most important information that the primary audience needs summarized in the first paragraph or section? (Q10)’, and ‘Does the material consistently explain the meaning of the numbers and units used? (Q16)’. With regard to Q10, usage precautions, such as advising immediate discontinuation of product usage and recommending consultation with a doctor if any physical changes are noticed, were described at the bottom of the label without any particular emphasis. For Q7, certain sentences indicated by the Consumer Affairs Agency (CAA) included technical jargon that was not commonly used by the public. The results indicated that the readability and location of the main message, in particular, should be improved.

### Evaluation of FFC labelling through user-testing

In addition to evaluation from the providers’ perspective, it is imperative to assess FFC labelling from the end-user’s standpoint. Therefore, a comprehensive evaluation from both perspectives is crucial.

User testing is used to assess whether users can easily access and understand FFC[24,25]. It is widely employed for assessing the efficacy of consumer health information, ranging from booklets and leaflets to online resources. User testing aims to enhance the understanding of provided information for consumers and patients [24–28]. The interviewer asked participants to answer questions about the content of the materials. When conducting user testing, it is recommended to employ a cohort of 10 participants at a time. This approach is well-established and supported by EU and Australian guidelines and meeting user testing criteria is one of the conditions for the approval of new medicines in the EU [26–28]. This methodology has been widely used, as demonstrated by Raynor DK et al. [29–31].

Our initial user testing in Japan targeted the Drug Guides for Patients, which are the label information of prescription drugs for patients [32]. Subsequently, we have continued user testing and gained experience in this field [33]. In the test, the passing criterion is as follows: 90% of the participants should successfully locate and 90% should be able to understand the information.

In this study, we evaluated five FFC labels by the F-CCI and conducted user testing on five FFC (Fig 2.). Interviews with 50 participants (five cohorts of 10 participants) in the user testing were conducted to gain insights into users’ attitudes to enhance the overall quality of FFC labelling.

**Fig 2.**
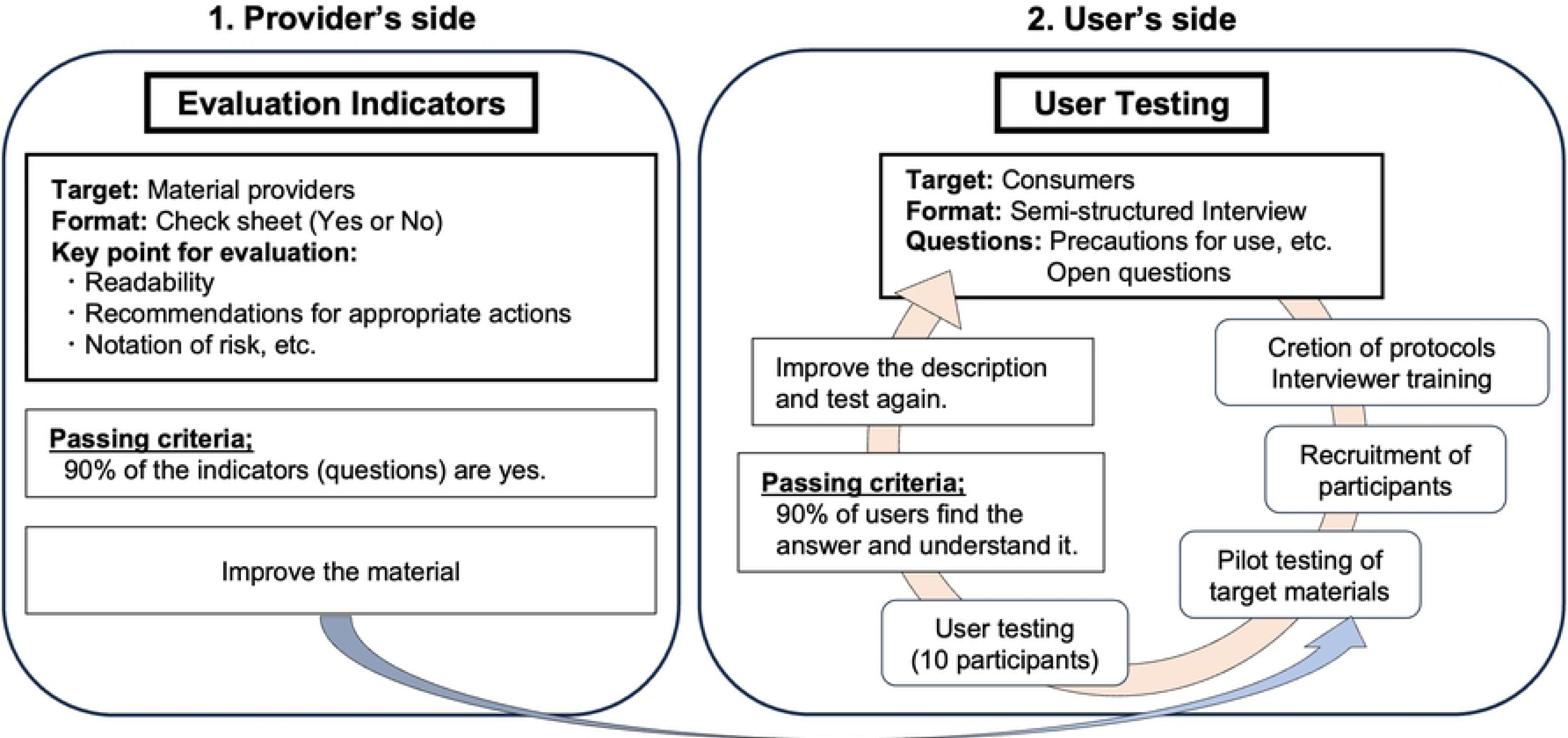
Evaluation of integrated usability of Health Information Materials.

## MATERIALS AND METHODS

### Materials

On the FFC search site provided by the CAA [17], we searched for FFC relevant to keywords ‘triglyceride’, ‘presbyopia’, ‘absorption of sugar and fat’, ‘hypertension’, and ‘cholesterol’. These topics are of particular interest to middle-aged and older adults. After reviewing approximately 100 labelling of FFC in a preliminary study, we selected five products, each with distinct claims of functionality that were considered commonplace. Table 3 provides an overview of these five products. Subsequently, we purchased each product and evaluated its labelling content. The labelling and labelling sample (Form VI of the submitted claim) can be found on the CAA website (accessed on 5 January 2020).

**Table 3.** Overview of foods with function claims subjected to user testing.

|  | Functional substance | Sales copy | Submitted claim | Containers or packaging | Product format |
| --- | --- | --- | --- | --- | --- |
| Product A | Eicosapentaenoic acid, Docosahexaenoic acid: substance A | Reduces triglyceride in individuals with elevated triglyceride levels. | This product contains substance A, known for its triglyceride-reducing properties and reported health benefits for individuals with elevated triglycerides. | Cardboard box | Soft capsules |
| Product B | Lutein astaxanthin (as free form), Cyanidin-3-glucoside, Docosahexaenoic acid : substance B | Promotes improved focus on near objects to enhance eye health in middle-aged and older individuals.<br>Designed for alleviating difficulties reading small print up close, eliminating the need for reliance on glasses. | This product contains substance B, known to assist with near focus and alleviate neck and shoulder pain associated with eye strain. | Plastic pouch | Tablets |
| Product C | Non-digestible dextrin (dietary fibre): substance C | Reduces the absorption of dietary sugar and fat. | This product contains substance C, reported to suppress the absorption of dietary fat and sugar. | PET bottle | Liquid |
| Product D | Lactotripeptide (Valyl-Prolyl-Proline, Isoleucyl-Prolyl-Proline): substance D | Designed for individuals with high blood pressure. | This product contains substance D, reported to lower blood pressure in individuals with elevated levels. It is recommended specifically for those with high blood pressure. | PET bottle | Liquid |
| Product E | Pine bark-derived procyanidins (as procyanidin B1): substance E | Lowers bad cholesterol. Controls cholesterol (LDL). | This product contains substance E, reported to lower bad cholesterol (LDL) levels. Consequently, this beverage is recommended for individuals concerned about bad cholesterol (LDL). | PET bottle | Liquid |

PET: Polyethylene terephthalate, LDL: Low-density lipoprotein

### Participants

For generalizability of the outcomes of user testing, it is imperative to carefully recruit a subject sample that accurately reflects the characteristics of users of the specific product under consideration [26–28]. The distribution of variables such as age, gender, literacy level (e.g., education), and others within the subject sample should closely mirror the distribution observed among the actual users of the product in question. It is noteworthy, however, that the utilization of random sampling may not be necessary in all cases [28]. We conducted the recruitment between from July 1st 2020 to September 30th 2020 using recruitment flyer distribution, SNS and through a market research company.

### Criteria for Participant Suitability

The designated number of subjects per product is set at 10, given the execution of five cohorts, resulting in the recruitment of a total of 50 subjects. To guarantee a comprehensive representation within the target group, the following criteria are established for the inclusion of participants:

Age: Individuals aged between 30 and 70 years, aligning with the age range during which FFC products are most commonly utilized.

Gender: Each gender category must be represented by a minimum of four individuals.

Literacy Level: High school, and vocational school graduates or equivalents are to be included, ensuring diverse educational backgrounds within the target group.

### Occupation: Includes two or more people who do not regularly use written information as part of their occupation

#### Exclusion Criteria for Participants

1. Individuals who are currently using or have used FFC products under investigation within the past 6 months.
2. FFC is utilized by a family member residing in the same household.
3. Individuals involved in health professions, pharmaceutical professions, occupations associated with health products, or those with prior work experience in these domains.
4. P articipants who have been subjects of a user test within six months.

Taking into account a balanced distribution in terms of sex, age, and literacy level (education background), as described in Table 4 [26,27]. We provided potential participants with written explanations outlining the purpose and methods of the test and obtained their written informed consent. No people refused to participate or dropped out of the user-test.

**Table 4.**
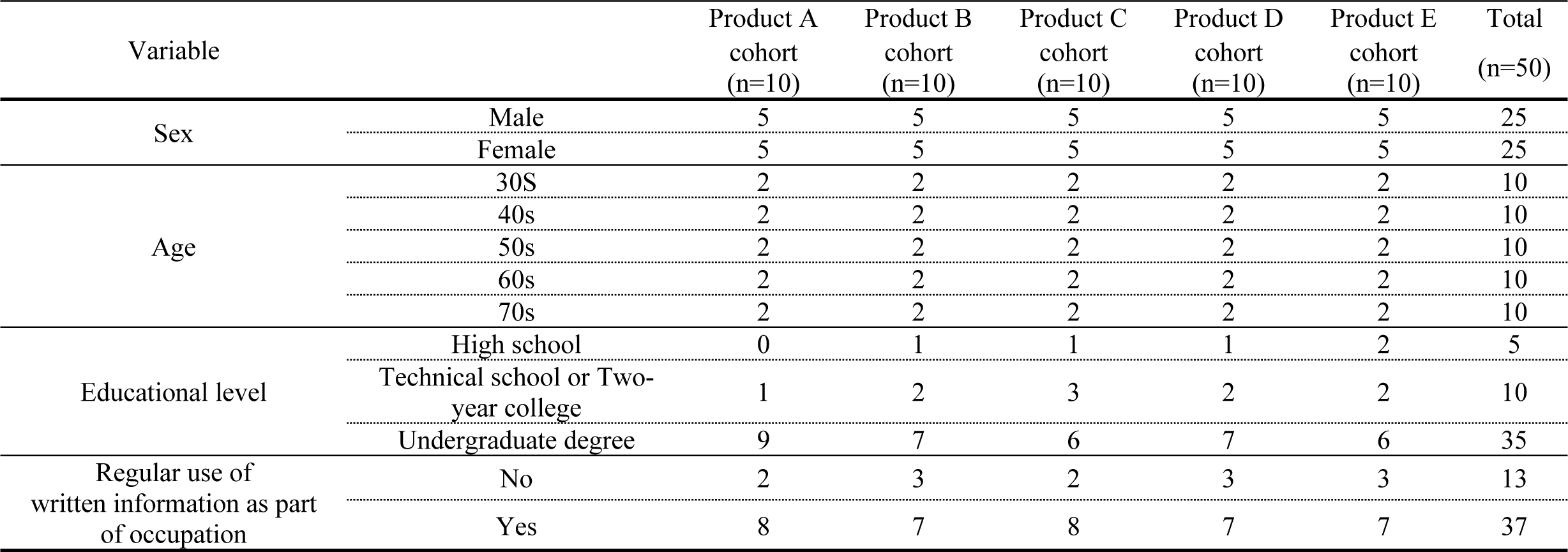
Characteristics of participants of the user testing of foods with function claims.

#### User-Testing Procedure

The user-testing was conducted as follows:

(1) Preliminary preparation
  i) Development of protocols [34]

The user testing procedures and methods were consolidated into a protocol. Specific questions were developed for products.

ii) Interviewers

The two interviewers underwent training to standardise their levels of observational and listening skills before engaging in user-testing. They are university employees with Ph.D. They are qualified interviewers accredited by the Japanese Interviewer Association.

iii) Conducting a pilot test

The user-testing of the pilot test was conducted from October 1st 2020 to January 31th, 2021 including. We conducted a pilot test with three participants to assess the appropriateness of the user-testing procedure, the manner and wording of the questions, and response time settings.

Subsequently, the protocol was adjusted based on the findings. Written informed consent was obtained from those participants.

(2) User-testing

The user-testing took place between April 1st 2021 and December 30th 2022. Written informed consent was obtained from all subjects involved in the study.

i) Place and timing of the interview

A quiet room with adequate privacy was prepared for the participants to relax and be interviewed at home or our work place. Each interview was scheduled to last approximately 1 h, including the time needed to explain the user testing procedure and obtain consent. The interviews were recorded with the participants’ consent.

ii) User testing questions

We developed a dozen questions on labelling according to the characteristics of each of the five FFC labels. Among them, a total of 10 questions were selected. The order of them was arranged randomly rather than following the order on the label. These questions were short and open-ended, as outlined in Table 5. Standardised questions were prepared addressing the appropriate and safe use of the five products. Finally, participants were asked to provide feedback on the comprehensibility, issues, design, and layout of the labels (Table 6).

**Table 5.** Questions on the content of foods with function claims labelling in the user test.

|  |  |
| --- | --- |
| Q1 | What should you pay attention to in your diet? |
| Q2 | What should you do if you are taking medicines? |
| Q3 | What is the recommended daily intake? |
| Q4 | Who is not subject to the development of this product? |
| Q5 | If you are ill, what should you do? |
| Q6 | If you have allergies, where on the label can you find the relevant information? |
| Q7 | What should you do if you experience an unpleasant reaction or develop a concerning symptom? |
| Q8 | What precautions should be observed when storing this product? |
| Q9 | What considerations should be kept in mind when handling this product? |
| Q10 | Where can you find the contact details of the food business operator? |

**Table 6.**
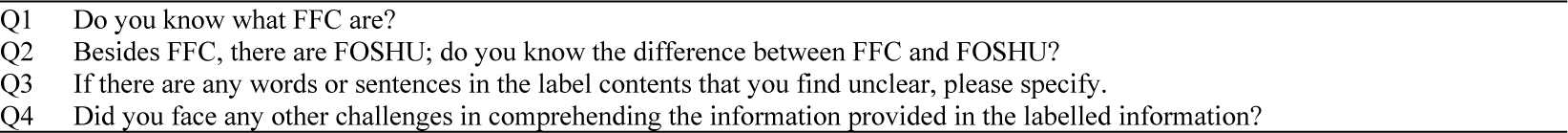
Questions on labelling foods with functional claims for participants’ comments.

|  |  |
| --- | --- |
| Q1 | Do you know what FFC are? |
| Q2 | Besides FFC, there are FOSHU; do you know the difference between FFC and FOSHU? |
| Q3 | If there are any words or sentences in the label contents that you find unclear, please specify. |
| Q4 | Did you face any other challenges in comprehending the information provided in the labelled information? |

### Outcome Measurement

The participants were asked to locate the relevant information, and the response time was recorded for each question. In addition to providing answers, participants were asked to rephrase the information in their own words to assess their understanding of the materials. Two cut-off points, at 1 and 2 min of response time, were used to evaluate participants’ understanding. The 1-min cut-off was established based on our previous user testing on drug information, which indicated that, on average, people need 1 min to understand every 1,000 characters of information correctly [32,33]. Given that FFC labelling contains ≤ 1,000 characters, in theory, 1 min should suffice. The 2-min cut-off was also used based on the results from the pilot test, indicating that participants needed approximately 2 min to answer each FFC-related question. The product was considered to pass the test if 90% or more of the participants could find and correctly understand the information for all 10 questions before the specified cut-off time. If a participant could not find the answer within 2 min, their response time was recorded as 2 min.

### Qualitative Analysis of Participants’ Comments

Using semi-structured interviews, participants were asked to respond to the questions following each user test (Table 6). In our analysis, we incorporated elements of the KJ Method, a qualitative research strategy developed by Kawakita [35,36]. Qualitative analysis was conducted for each question to gain insights from participants’ responses.

### Comparison between the current version and the revised version of the standardised wording

Our analysis using the F-CCI indicated potential areas for improvement in the standardised wording included in the FFC label, originally developed by CAA. Some of the wordings were considered difficult to understand. Furthermore, the user testing conducted in this study showed that the current wording was difficult to understand and time-consuming. Therefore, we developed a revised version of the standardised wording to be used on the FFC label and compared it against the current version developed by CAA (Fig 3). To enhance the comprehensibility of container and packaging labels, a QR code can be added to the label. This code can direct the user to a page with clear and concise explanations. The terms ’Submitted Claim’ and ’Individual Review’, as well as the distinction between FFC and Food for Specified Health Uses (FOSHU), were explained.

**Fig 3.**
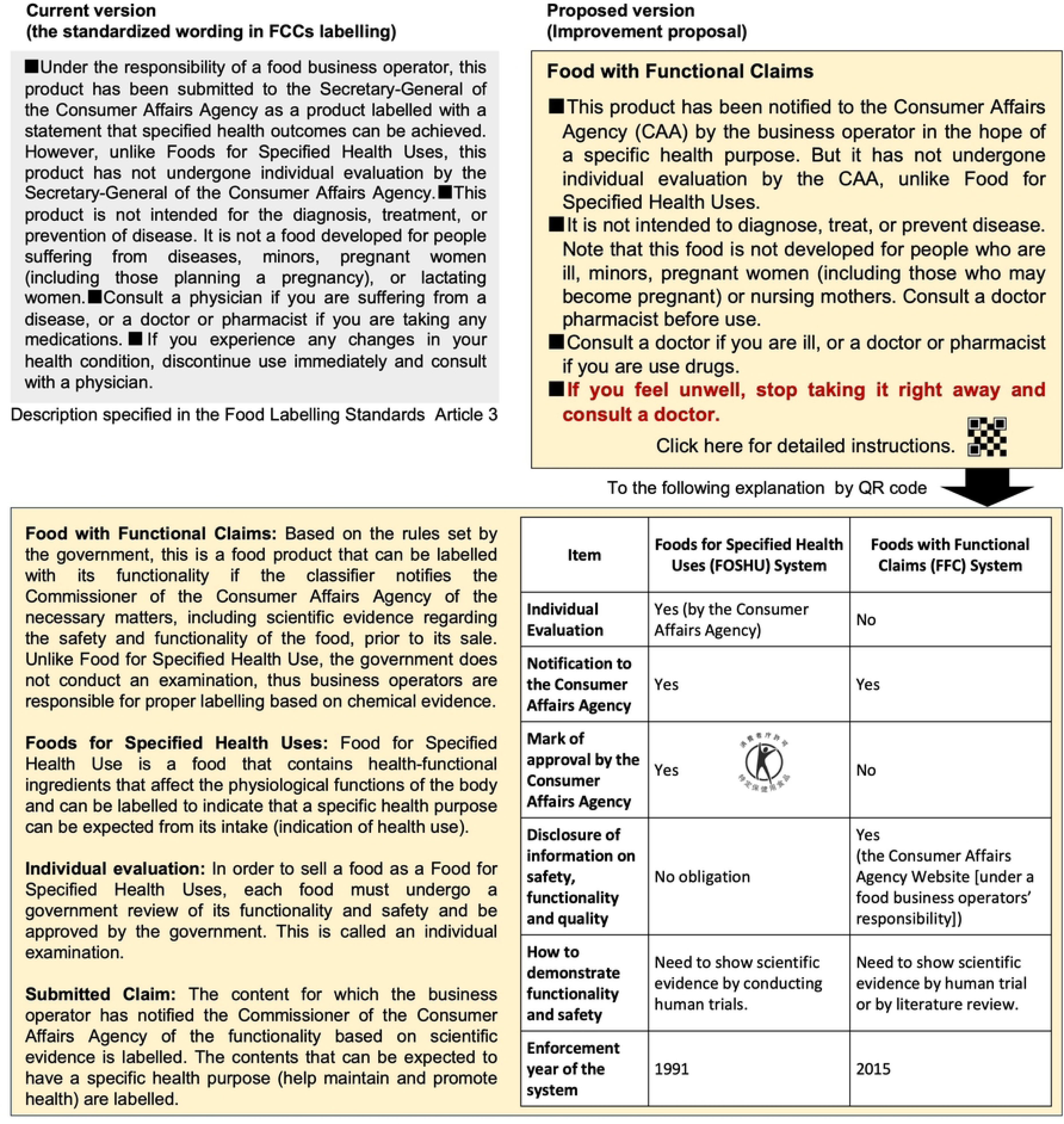
The description is specified in the Food Labelling Standards and the proposed revision.

Participants assessed the current and revised versions on a 5-point scale (5 = very easy to understand, 4 = easy to understand, 3 = neither, 2 = difficult to understand, and 1 = very difficult to understand) across four questions (Q1: Size, legibility, and length; Q2: terms and sentences; Q3: Usefulness of the information; Q4: Overall evaluation). To compare participant evaluations of the current and the revised versions, we conducted a paired t-test with an alpha of 0.05. This analysis was performed using SPSS Statistics Version 29.0.1.0 (IBM, Armonk, NY, USA).

## RESULTS

### Accessibility and understandability of the FFC labelling in user-testing

When the 2-min cut-off was used, only one product (product B) among the five user-tested products met the threshold of 90% for all 10 questions (Table 7-1). However, the overall results were relatively positive. Products A and E achieved 90% or more correct responses for all questions but one (Q6). Product C missed the passing score for Q4 and Q5. Product D, whose labelling had the smallest font size, demonstrated the poorest performance of the five products, with two questions (Q1, Q2) failing to meet the criterion. Questions 3 to 10 had high percentages of 90% or more correct responses for all products, whereas a question on concomitant medications (Q2) had the lowest percentages of correct responses across all five products, ranging from 70% to 100%. When participants were able to find an answer to a question, it was considered they understood its content.

**Table 7-1.**
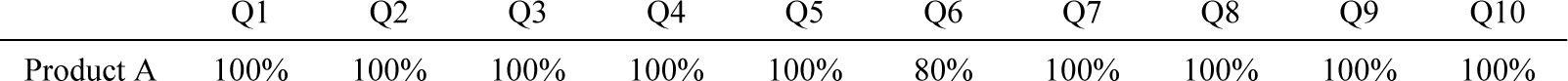
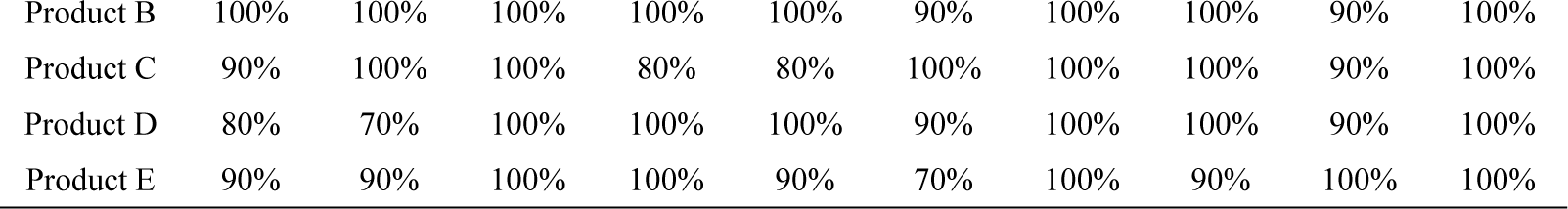
The proportion of participants who correctly identified answers for each question within 2 min.

|  | Q1 | Q2 | Q3 | Q4 | Q5 | Q6 | Q7 | Q8 | Q9 | Q10 |
| --- | --- | --- | --- | --- | --- | --- | --- | --- | --- | --- |
| Product A | 100% | 100% | 100% | 100% | 100% | 80% | 100% | 100% | 100% | 100% |
| Product B | 100% | 100% | 100% | 100% | 100% | 90% | 100% | 100% | 90% | 100% |
| Product C | 90% | 100% | 100% | 80% | 80% | 100% | 100% | 100% | 90% | 100% |
| Product D | 80% | 70% | 100% | 100% | 100% | 90% | 100% | 100% | 90% | 100% |
| Product E | 90% | 90% | 100% | 100% | 90% | 70% | 100% | 90% | 100% | 100% |

When evaluated at 1 min, the overall performance was considerably poorer than that at 2 min, with no product meeting the 90% criterion (Table 7-2). Product A and B were still satisfactory-performing products, missing the passing score for two questions (Q1, Q6). Product E missed the passing score of four questions (Q2, Q6, Q7, Q8). Product D remained the worst-performing product, not meeting the 90% threshold for more than half of the questions. The proportion of correct answers within 1 min remained the lowest for Q6 (50–90%), with lower proportions for Q1 (50–90%), Q2 (50–100%), Q8 (30–100%), and Q9 (30–100%), whereas all products successfully passed Q3 and Q10 in less than 1 min. The median access times for each product and question are listed in S1Table.

**Table 7-2.** The proportion of participants who correctly identified answers for each question within 1 min.

|  | Q1 | Q2 | Q3 | Q4 | Q5 | Q6 | Q7 | Q8 | Q9 | Q10 |
| --- | --- | --- | --- | --- | --- | --- | --- | --- | --- | --- |
| Product A | 80% | 100% | 100% | 100% | 90% | 50% | 100% | 100% | 100% | 100% |
| Product B | 80% | 90% | 100% | 100% | 100% | 80% | 100% | 90% | 90% | 90% |
| Product C | 90% | 70% | 90% | 70% | 60% | 90% | 80% | 30% | 50% | 90% |
| Product D | 50% | 50% | 100% | 100% | 50% | 80% | 100% | 80% | 30% | 90% |
| Product E | 90% | 70% | 100% | 100% | 90% | 70% | 80% | 80% | 100% | 100% |

### Qualitative analysis of participants’ comments

For each question, comments were collected during the interviews, and similar comments were grouped (Table 8). Many participants had insufficient knowledge of FFC and perceived it as potentially beneficial for their health based on its image. Additionally, a significant number were unaware of the distinction between FFC and FOSHU, with 30% incorrectly believing that FFC was superior in effectiveness to FOSHU. The labelling included numerous technical terms such as “Submitted Claim” and “Indivisual Review”. Some sentences posed challenges in comprehension and interpretation. For instance, it was difficult to discern the intended meaning of the statement “This product is not a food developed for people suffering from diseases, minors, pregnant women (including those planning a pregnancy), or lactating women.” As a result, the participants were unclear as to whether the relevant individuals were allowed to take it or not.

**Table 8.**
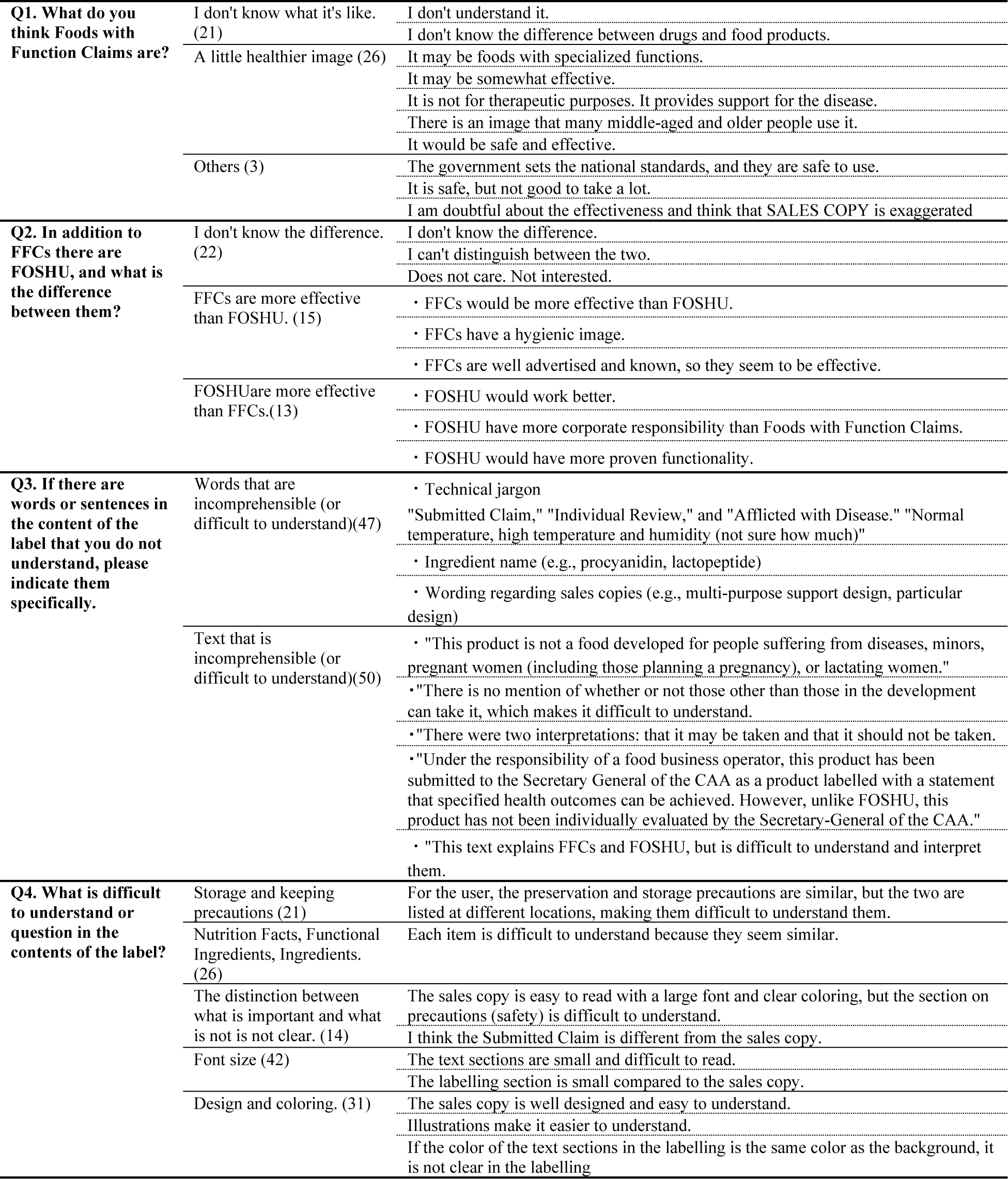
Classification of participants’ comments on the labelling of FFC.

While the sales copy was easily understood because of its large font size and good design, the labelling itself presented difficulties in reading due to the small font size and the shape of its container or packaging.

### Comparison between the current version and the revised version of the standardised wording

In this user-test of FFC, the participants’ comments showed that the standardised wording developed by CAA (current version) is difficult to understand, consistent with our previous finding. Therefore, we developed a proposed revision to improve the original version developed by CAA and conducted additional user testing to compare the two versions (Fig 3). “Due to limited space on the packaging, a QR code is provided for accessing more detailed information, including terminology, which can be obtained by scanning the QR code. 20 participants were asked to compare them on a five-point scale for product B. The results of the evaluation are shown in Table 9. The revised version received higher ratings than the current version across all four elements (Size, legibility, and length; terms and sentences; usefulness of the information; and overall evaluation). The differences were all statistically significant.

**Table 9.** Comparison between the current version and the revised version of the standardised wording.

| Question | Current version | Revised version | P-value |
| --- | --- | --- | --- |
| Q1 Size, legibility, and length | 3.1 ± 0.7 | 4.3 ± 0.6 | .005 |
| Q2 Terms and sentences | 2.7 ± 0.6 | 4.3 ± 0.8 | .001 |
| Q3 Usefulness of the information | 2.9 ± 0.5 | 4.3 ± 0.6 | <.001 |
| Q4 Overall evaluation | 2.6 ± 0.7 | 4.6 ± 0.5 | <.001 |

N=20

Current version: the standardised wording shown by the CAA in labelling Revised version: Improvement Proposal based on the current version

## DISCUSSION

To date, there has been a dearth of studies scrutinizing health food labels from both provider and user perspectives, even in international contexts. In this study, we conducted user testing and evaluated five FFC, which we had previously evaluated using the F-CCI. The results of this user test showed that, in many cases, FFC labels provide inadequate explanations and are difficult to understand. In consumer research on food labelling, qualitative research should be conducted from the consumer’s point of view, using a range of approaches including observation and semi-directive interviews in addition to questionnaire surveys [37]. This study marks the first user testing of FFC labelling, complemented by qualitative research, providing novel insights into the evaluation of FFC labelling. When assessing the labelling of FFC, it is crucial to approach the evaluation from the perspectives of both the provider and the end-user.

We previously evaluated FFC labelling from the provider’s perspective [12]. None of the five products met the acceptance criteria for the questions of F-CCI. Certain sentences indicated by the CAA included technical jargon that was not commonly used by the public.

Important information was often not immediately accessible because it was not summarized in the first section or was scattered throughout the label. Based on our previous user testing on drug-related information and considering the word count of the FFC label, we initially expected that one minute would suffice for consumers to capture and comprehend the information accurately. However, the testing revealed consumers needed 2 minutes, twice as long as for drug-related information. One factor contributing to the longer time required for response is that the order of entry, position of entry, and font size in the FFC label are not specified, unlike those in drug labels. This underscores a clear need for improvements in FFC labelling to enhance information accessibility and consumer understanding. In particular, precautions for safe use need to be more clearly alerted in view of the recent incident in Japan. It should also be noted that physical health care is particularly important in the case of self-care.

However, during the interviews, it became evident that the participants not only lacked sufficient knowledge about FFC, but also held misconceptions. This could pose a fundamental challenge to the proper use of FFC. Taking into account consumers’ health literacy levels, information providers should provide easy-to-understand information.

The evaluations of the two surveys yielded largely consistent outcomes, highlighting the need for improved labelling to enhance consumer safety and product usage. Overall, there is often a focus on product promotion through design and sales copy rather than facilitating consumer access to important information and comprehension of messages. In particular, precautions for safe use must be presented in a manner that is more easily understandable for consumers.

One limitation of this study was that it only evaluated five FFC. In selecting the five products, more than 100 FFC labelling were consulted, and the labelling showed a similar trend in labelling content, with the wording portion recommended by the CAA accounting for about half of the labelling. Nevertheless, valuable insights into consumer perceptions were garnered through live feedback from 50 interviewees. However, future research should explore a broader spectrum of FFC.

We believe that the standardized language used in food labelling requires improvement as it currently contains numerous technical terms that pose difficulties for consumers to comprehend. In the development of health information materials, such as FFC, the newly established system facilitates the creation of optimal materials. This is achieved by enabling providers to assess and enhance the materials using a communication index as a specified indicator, followed by a validation process to ascertain their effectiveness and assess consumer comprehension. In future, we intend to promote a website that we have developed to evaluate the usefulness of health-related information materials [34].

## CONCLUSIONS

In this study, we undertook the first user testing of FFC labelling in Japan to ascertain users’ perceptions and comprehension. The results indicated that consumers encountered challenges in locating and understanding information within the current FFC labelling. The evaluation of the user test underscores the need to improve the presentation of key information to ensure safe and appropriate use of FFC, given that most consumers are not familiar with FFC. Moreover, how information is currently provided on FFC labelling may not be adequate in certain situations.

In the development and application of FFC labelling for effective risk-benefit communication, a critical evaluation is imperative from both the provider’s and user’s viewpoints. The importance of establishing an integrated method for assessing usefulness becomes paramount in this context.

This initiative holds substantial significance as it has the potential to significantly contribute to consumer decision-making and the secure utilization of health food products including FFC.

## Author Contributions

Conceptualization, Michiko Yamamoto; Data curation, Ken Yamamoto and Michiko Yamamoto; Formal analysis, Ken Yamamoto and Rain Yamamoto; Funding acquisition, Michiko Yamamoto; Investigation, Michiko Yamamoto, Hiromi Takano-Ohmuro and Junji Saruwatari; Methodology, Michiko Yamamoto; Project administration, Michiko Yamamoto; Software, Ken Yamamoto, Michiko Yamamoto and Junji Saruwatari; Supervision, Michiko Yamamoto and Rain Yamamoto; Validation, Ken Yamamoto, Michiko Yamamoto, Hiromi Takano-Ohmuro, Rain Yamamoto and Junji Saruwatari; Visualization, Ken Yamamoto and Michiko Yamamoto; Writing – original draft, Ken Yamamoto and Michiko Yamamoto; Writing – review & editing, Ken Yamamoto, Michiko Yamamoto, Hiromi Takano-Ohmuro, Rain Yamamoto and Junji Saruwatari.

## Institutional Review Board Statement

The study was conducted according to the guidelines of the Declaration of Helsinki, and approved by the Institutional Review Board of Kumamoto Graduate School of Pharmaceutical Sciences (No.2039)

## Informed Consent Statement

Written informed consent was obtained from all subjects involved in the study.

## Data Availability Statement

The data that support the findings of this study are available on request from the corresponding author. The data are not publicly available due to privacy or ethical restrictions.

## Acknowledgments

We thank all the participants of the user tests.

## Supporting information

**S1 Fig. Types of food with health claims**

**S1 Table. Accessibility per question for each product during the user testing IQR: interquartile range**

